# Association of Human Cytomegalovirus exposure with tuberculosis disease in South African adults with presumptive tuberculosis

**DOI:** 10.64898/2026.02.02.26345431

**Authors:** Derrick Semugenze, Arthur Chiwaya, George William Kasule, James Sserubiri, Rose Nabatanzi, Byron W P Reeve, Zaida Palmer, Hridesh Mishra, Achilles Katamba, Alberto García-Basteiro, Moses L Joloba, Grant Theron, Frank Cobelens, Willy Ssengooba

**Affiliations:** Department of Immunology and Molecular Biology, Makerere University College of Health Sciences Kampala, Uganda; Department of Global Health and Amsterdam Institute for Global Health and Development, Amsterdam University Medical Centers Location University of Amsterdam, Amsterdam, The Netherlands; DSI-NRF Centre of Excellence for Biomedical Tuberculosis Research; South African Medical Research Council Centre for Tuberculosis Research; Division of Molecular Biology and Human Genetics, Faculty of Medicine and Health Sciences, Stellenbosch University, Cape Town, South Africa; Sandra A. Rotman (SAR) Laboratories, Sandra Rotman Centre for Global Health, University Health Network-Toronto General Hospital, Toronto, ON, Canada; Clinical Epidemiology & Biostatistics Unit, Department of Medicine, Makerere University College of Health Sciences, Kampala, Uganda; Centro de Investigação em Saúde de Manhiça (CISM), Maputo, Mozambique; ISGlobal, Barcelona Centre for International Health Research, Hospital Clínic - Universitat de Barcelona, Barcelona, Spain; Department of Medical Microbiology, Makerere University College of Health Sciences Kampala, Uganda

**Author notes:** Correspondence to: Dr. Willy Ssengooba, Department of Medical Microbiology, Makerere University College of Health Sciences Kampala, Uganda. Deceased (Byron W P Reeve).

**Keywords:** Cytomegalovirus, risk factors, tuberculosis, susceptibility, reactivation, reinfection

## Abstract

Recent studies suggested that human cytomegalovirus (HCMV) exposure may increase tuberculosis (TB) disease risk. We assessed the association between active HCMV infection and recent HCMV exposure with tuberculosis (TB) disease among TB-presumptive South African adults. This was a nested case-control analysis that utilized stored plasma and serum samples collected from adults (≥18 years) with presumptive TB self-presenting to primary care clinics in in the Kraaifontein District in Cape Town, South Africa. Cases (n=98) and HIV status frequency matched controls (n=199) basing on mycobacterial culture and or GeneXpert Ultra were included in the study. HCMV DNAemia was detected by qPCR well as current HCMV reactivation or reinfection and recent HCMV infection, reactivation or reinfection were categorized using PCR and serology (IgM and IgG avidity ELISA) results. The median age of all participants was 37 years (IQR 29-47), 164 (55.2%) were male and 119 (40.1%) had previous TB treatment. Overall, 21 (7.1%) had HCMV DNAemia, 19 (6.4%) had positive HCMV IgM and 2 (0.7%) had low HCMV avidity. In a logistic regression model adjusting for age, gender, HIV status and BMI, TB disease was associated with current HCMV reactivation or reinfection [adjusted odds ratio (aOR) 4.88, 95%CI 1.59-16.31, p=0.007]. There was no association with recent HCMV infection, reactivation or reinfection. Unlike recent HCMV infection, reactivation or reinfection, active HCMV replication although not frequent was associated with TB disease which suggests that TB disease or an underlying common factor reactivates HCMV replication in this population.

## Introduction

About 10.8 million people developed tuberculosis (TB) disease in 2023 and the number of cases have been reported to be increasing every single year since the COVID-19 pandemic(1). About 5 - 15% of the *Mycobacterium tuberculosis* (Mtb) latently infected individuals progress to active disease within their lifetime(2), but knowledge on the factors that lead to disease progression is incomplete. Exposure to human cytomegalovirus (HCMV), a herpesvirus that remains present lifelong after (re-)infection and can reactivate, has been suggested as one of such factors(3). In a recent systematic review, all studies that investigated HCMV associated it with TB disease(4). HCMV infection was unrelated to tuberculin skin test conversion(5), suggesting this association may reflect enhanced disease progression due to active HCMV infection. Between 60% and 90% of adults are estimated to be HCMV seropositive worldwide with highest prevalence in non-Caucasian populations and low socioeconomic settings(6). HCMV modulates the host immunity, amongst others by causing change overtime in the T-cell repertoire through significant depletion of naïve T-cells and increased numbers of memory T cells, a phenomenon referred to as immunosenescence(7). This might possibly reduce the ability of the host to mount an effective adaptive immune response against pathogens including Mtb; since T-cells are crucial for controlling Mtb infection this could lead to unstable control of Mtb and disease progression(8). While other studies have looked at circulating anti-HCMV IgG levels, two studies in South Africa showed increased risk of TB disease among infants and children with HCMV shedding and HCMV-specific interferon-gamma responses, respectively (5, 9). These studies followed up infants from birth, and hence associated primary HCMV infection with primary TB disease (5). Whether these findings can be extrapolated to adults is unknown. Adults usually have reactivations and reinfections of HCMV rather than primary infections (10) and in TB endemic countries, adults are likely to have reactivation TB or even reinfection TB rather than primary disease (11). We therefore set out to assess whether HCMV viremia and reactivation or reinfection is associated with TB disease in South African TB presumptive adults.

## Materials and methods

### Study design and participant flow

Adults (≥ 18 years) with presumptive pulmonary TB disease(12), not currently on treatment or within the previous two months were consecutively recruited between 06^th^ February 2016 and 22^nd^ February 2023 at Primary health care clinics in the Kraaifontein District in Cape Town, South Africa (Figure I). De-identified demographic, clinical and laboratory data relevant to this study that was captured in REDCap(13) was downloaded on 12^th^ June 2024. Participants with non-actionable culture results, those with missing, inappropriate or with missing sample volumes were excluded from subsequent analysis. Tuberculosis cases [culture positive and or GeneXpert MTB/RIF Ultra (Cepheid Sunnyvale, CA, United States) positive] and controls who were frequency matched by HIV status were randomly selected from the eligible participants. Both cases and controls were recruited from the Primary health care clinics.

**Figure I.**
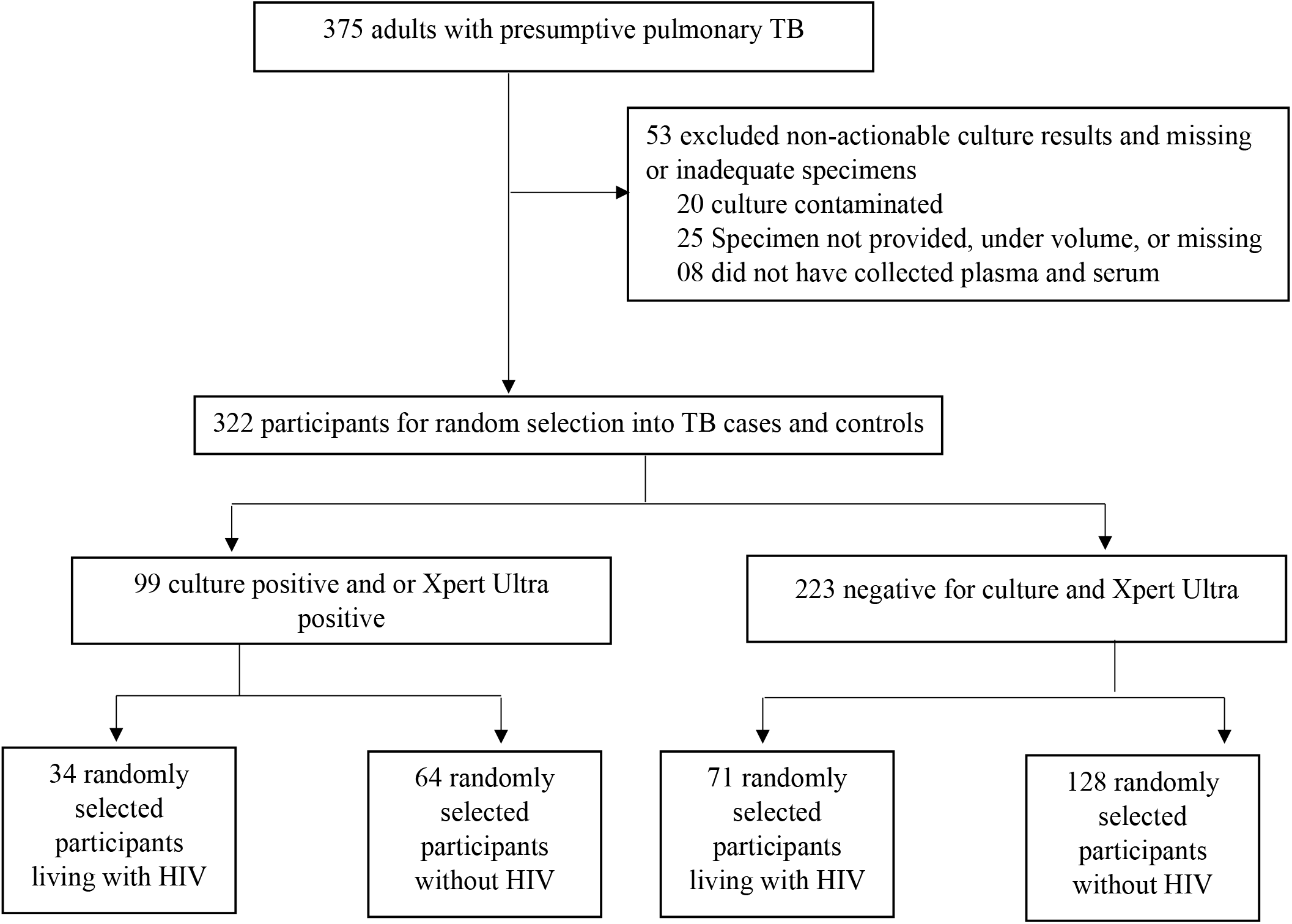
Flowchart illustrating the selection and categorization process of study participants. TB: tuberculosis

### Laboratory procedures

Plasma and serum samples were initially stored at -80°C and shipped on dry ice to the Integrated Biorepository of H3Africa Uganda, Makerere University, where they were immediately stored at -80°C before the analysis.

#### Real-time qPCR

HCMV DNAemia was determined by extracting viral DNA using QIAamp DSP Virus Spin kit (Qiagen, Hilden, Germany) following the manufacturer’s instructions. Real-time qPCR was done using artus CMV RG PCR kit (Qiagen, Hilden, Germany) on the QuantStudio5 platform using the following parameters: hot start enzyme activation at 95°C for 10 minutes, denaturation at 95°C for 15 seconds, annealing at 65°C for 30 seconds, and extension at 72°C for 20 seconds. Forty-five cycles were done during denaturation and annealing. This assay had a HCMV DNA limit of Detection (LoD) of 11.42 copies/ml in plasma.

#### Detection of HMCV specific IgM antibodies

Euroimmun HCMV ELISA Kits (Lübeck, Germany) with coated recombinant EI-p52 antigen were used according to the manufacturer’s instructions. The participant sera were diluted 1:101 before pipetting 100μl into the microtiter wells and incubated for 30 minutes. The wells were then washed. After that, 100μl of horseradish peroxidase-conjugated anti-human IgM antibodies were transferred into microtiter wells and still incubated for 30 minutes. The wells were then washed to remove the unbounded conjugate from immune complexes. The wells were incubated with TMB substrate for 15 minutes. The positive control was monitored for development of a blue color. 100μl of the stopping solution (sulfuric acid) was then added to stop the enzymatic reaction (the blue color turned to yellow). For measuring the optical density (OD) of the samples, the Biotek Synergy H1 Multi-Mode Microplate reader (Winooski, Vermont, USA) was used at 450 nm using 620nm as reference. Participants were considered seropositive when the ratio of extinction of the patient sample to that of the calibrator was ≥1.1.

#### Determination of HMCV specific IgG avidity antibodies

The manufacturer’s procedure was followed, similar to the one described above for IgM antibody determination, but with running each sample in duplicate and an additional step of adding 100μl of Urea and Phosphate Buffered Saline (PBS) in each duplicate followed by incubation at room temperature for 10 minutes after the step of washing out the diluted sera from the microtiter wells. Participants were considered to have low and high avidity antibodies when the relative avidity index (RAI) was <40% and >60% respectively.

### Statistical analysis

We used R studio software (Version R.4.5.1) for data analysis. Sociodemographic and clinical characteristics of the participants were summarized using median for age and frequency tables for other variables. We determined crude odds ratios and then performed univariate logistic regression to assess the association between each independent variable and TB disease. A Directed Acyclic Graph was drawn (Supplementary figure 1) to categorize different variables in the dataset into potential confounders, mediators and effect modifiers for the association between HCMV DNAemia, primary infection or reactivation/reinfection and TB disease based on the scientific knowledge and published literature, as guided by(14).

Our primary analysis investigated the association of TB disease with HCMV DNAemia. Potential confounders were included in multivariable logistic regression models along with age, gender and HIV status if they showed a difference of ≥10% between the crude odds ratio and adjusted odds ratio, a restriction added to prevent overfitting. Body Mass Index (BMI) was also examined for effect modification of the association between HCMV DNAemia and TB disease. All analyses used a p=0.05 significance level. Hemoglobin attenuated the association after including it in the multivariable logistic regression model which prompted its examination in a multivariable mediation analysis estimating direct, indirect and total effect with nonparametric bootstrap confidence intervals (1000 simulations) using the mediation R package.

Our secondary analysis investigated the association of TB disease with past HCMV infection, primary HCMV infection, current and recent HCMV reactivation or reinfection, for which four categories were created (Box).

**Box: Categories of HCMV exposure applied in secondary analysis**

**Primary HCMV infection**: *Participants with low anti-HCMV IgG avidity* irrespective of *HCMV DNAemia and anti-HCMV IgM results*

**Current HCMV reactivation or reinfection**: *Participants with HCMV DNAemia and high anti-HCMV IgG avidity irrespective of anti-HCMV IgM results*

**Recent HCMV reactivation or reinfection**: *Participants without HCMV DNAemia, and positive for anti-HCMV IgM with high anti-HCMV IgG avidity*

**Past HCMV infection**: *Participants without HCMV DNAemia who tested negative for anti-HCMV IgM and had high anti-HCMV IgG avidity and had high anti-HCMV IgG avidity*

The same procedure described above for HCMV DNAemia was used for identification of potential confounders and inclusion in the multivariable model.

Descriptive statistics were also conducted to assess the effect of HCMV viral load on several TB-related outcomes, including Time to TB Positivity (TTP) in mycobacterial culture, Mtb culture results, and variation according to previous TB history. Finally, TB disease classification was examined by categorizing individuals into those with positive TB cultures without prior TB and those with positive cultures reporting previous TB.

### Ethical considerations

The mother study (BAR-TB Dx) had obtained Ethics approval (Ethics Reference No: N14/10/136) and all participants had provided written informed consent for their stored samples to be used for future analyses. We obtained ethical approval from the Makerere University College of Health Sciences School of Biomedical Sciences Research and Ethics Committee (Approval number: SBS-2023-402) and Uganda National Council for Science and Technology (Approval number: HS3255ES).

## Results

### Description of study participants and characteristics of TB cases and controls

A total of 375 participants were screened to participate in this study. After the exclusion of 53 participants due to non-actionable culture results, missing samples, and collected samples with insufficient volumes, 322 were eligible to participate in the study. A total of 297 participants including 98 TB cases (34 PLWHIV and 64 non-HIV infected) and 199 controls (71 PLWHIV and 128 non-HIV infected) were selected for this study (Figure I).

The median age of all the participants was 37 years (interquartile range, IQR 29-47; Table I). The majority were of mixed race (194, 65.3%), between 18-35 years old (126, 42.4%) and male (164, 55.2%). All were symptomatic with cough reported by 292 (98.3%); 185 (62.3%) participants were current smokers and 199 (40.1%) reported previous TB treatment. The overall HIV prevalence was 297 (35.4%); 44 (44.9%) of the cases were underweight compared to 43 (21.6%) of the controls, and six (6.1%) were overweight as determined by BMI compared to 55 (27.6%) of the controls. The proportion of PLWHIV with CD4 counts <200 cells/μl was similar between cases (10.2%) and controls (8.0%). Among the cases, 12 were HCMV DNAemia positive (12.2%), six were anti-HCMV IgM positive (6.1%) and one had low anti-HCMV avidity (1.0%) (Table II).

**Table I:**
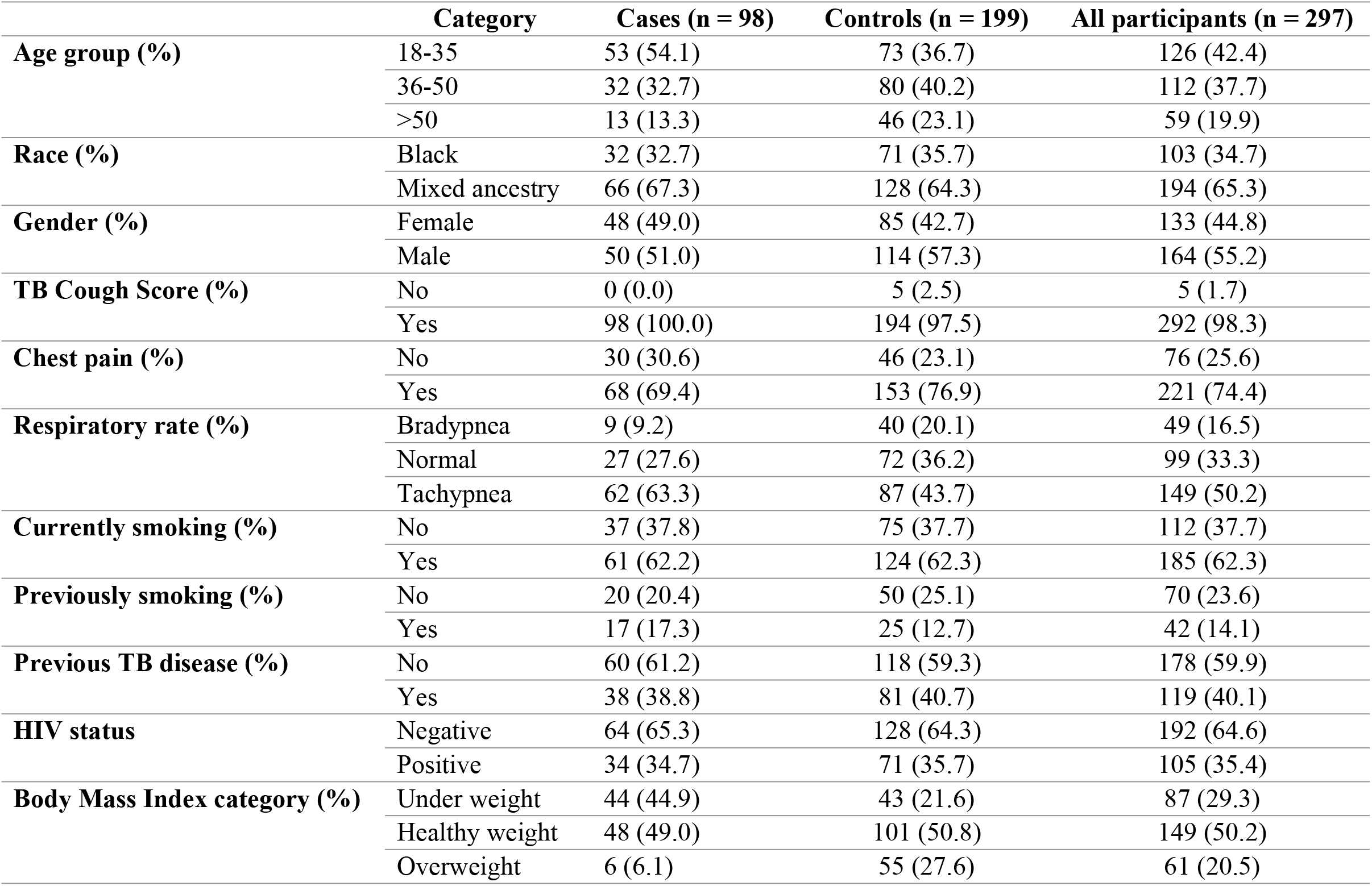
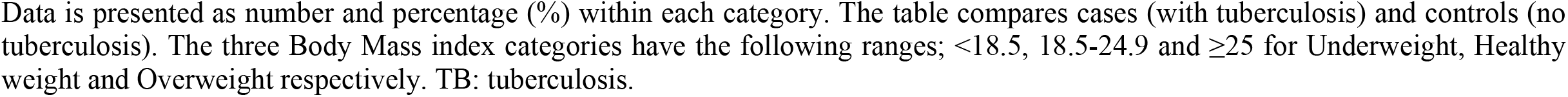
Demographic and clinical characteristics of study participants stratified by tuberculosis status.

|  | Category | Cases (n = 98) | Controls (n = 199) | All participants (n = 297) |
| --- | --- | --- | --- | --- |
| **Age group (%)** | 18-35 | 53 (54.1) | 73 (36.7) | 126 (42.4) |
|  | 36-50 | 32 (32.7) | 80 (40.2) | 112 (37.7) |
|  | >50 | 13 (13.3) | 46 (23.1) | 59 (19.9) |
| **Race (%)** | Black | 32 (32.7) | 71 (35.7) | 103 (34.7) |
|  | Mixed ancestry | 66 (67.3) | 128 (64.3) | 194 (65.3) |
| **Gender (%)** | Female | 48 (49.0) | 85 (42.7) | 133 (44.8) |
|  | Male | 50 (51.0) | 114 (57.3) | 164 (55.2) |
| **TB Cough Score (%)** | No | 0 (0.0) | 5 (2.5) | 5 (1.7) |
|  | Yes | 98 (100.0) | 194 (97.5) | 292 (98.3) |
| **Chest pain (%)** | No | 30 (30.6) | 46 (23.1) | 76 (25.6) |
|  | Yes | 68 (69.4) | 153 (76.9) | 221 (74.4) |
| **Respiratory rate (%)** | Bradypnea | 9 (9.2) | 40 (20.1) | 49 (16.5) |
|  | Normal | 27 (27.6) | 72 (36.2) | 99 (33.3) |
|  | Tachypnea | 62 (63.3) | 87 (43.7) | 149 (50.2) |
| **Currently smoking (%)** | No | 37 (37.8) | 75 (37.7) | 112 (37.7) |
|  | Yes | 61 (62.2) | 124 (62.3) | 185 (62.3) |
| **Previously smoking (%)** | No | 20 (20.4) | 50 (25.1) | 70 (23.6) |
|  | Yes | 17 (17.3) | 25 (12.7) | 42 (14.1) |
| **Previous TB disease (%)** | No | 60 (61.2) | 118 (59.3) | 178 (59.9) |
|  | Yes | 38 (38.8) | 81 (40.7) | 119 (40.1) |
| **HIV status** | Negative | 64 (65.3) | 128 (64.3) | 192 (64.6) |
|  | Positive | 34 (34.7) | 71 (35.7) | 105 (35.4) |
| **Body Mass Index category (%)** | Under weight | 44 (44.9) | 43 (21.6) | 87 (29.3) |
|  | Healthy weight | 48 (49.0) | 101 (50.8) | 149 (50.2) |
|  | Overweight | 6 (6.1) | 55 (27.6) | 61 (20.5) |

Data is presented as number and percentage (%) within each category. The table compares cases (with tuberculosis) and controls (no tuberculosis). The three Body Mass index categories have the following ranges; $<18.5$ , $18.5-24.9$ and $\geq 25$ for Underweight, Healthy weight and Overweight respectively. TB: tuberculosis.

When hemoglobin was included in the multivariable model as a covariate together with age, gender and HIV status, the association between TB status and HCMV DNAemia was attenuated and became statistically non-significant (aOR: 2.68, 95%CI 0.93–8.15; p=0.073). Hence performing a mediation analysis where we found that hemoglobin partially mediated the effect of HCMV DNAemia on TB disease, accounting for approximately one-fifth of the total effect (proportion mediated 0.21, p=0.028), with a statistically significant mediated effect (Average Causal Mediation Effect [ACME] 0.069, p=0.028) alongside a statistically significant direct effect (Average Direct Effect [ADE] 0.26, p=0.010; Supplementary table 2).

**Table II:** Laboratory investigation results of study participants stratified by tuberculosis status.

|  | **Category** | **Cases (n = 98)** | **Controls (n = 199)** | **All participants(n = 297)** |
| --- | --- | --- | --- | --- |
| **White Blood Cell count category (%)** | Leukocytosis | 32 (32.7) | 26 (13.1) | 58 (19.5) |
|  | Leukopenia | 7 (7.1) | 24 (12.1) | 31 (10.4) |
|  | Normal | 58 (59.2) | 145 (72.9) | 203 (68.4) |
| **Hemoglobin category (%)** | Anemic | 58 (59.2) | 60 (30.2) | 118 (39.7) |
|  | Normal | 40 (40.8) | 139 (69.8) | 179 (60.3) |
| **Platelet count category (%)** | Normal | 35 (35.7) | 155 (77.9) | 190 (64.0) |
|  | Thrombocytopenia | 3 (3.1) | 6 (3.0) | 9 (3.0) |
|  | Thrombocytosis | 59 (60.2) | 34 (17.1) | 93 (31.3) |
| **GeneXpert Ultra resistance (%)** | Indeterminate | 2 (2.0) | 0 (0.0) | 2 (0.7) |
|  | Rif resistance detected | 10 (10.2) | 0 (0.0) | 10 (3.4) |
|  | Rif resistance not detected | 86 (87.8) | 1 (0.5) | 87 (29.3) |
| **CD4 count (%)** | <200 cells/μl | 10 (10.2) | 16 (8.0) | 26 (8.8) |
| | >200 cells/μl | 23 (23.5) | 51 (25.6) | 74 (24.9) |
| **HCMV DNAemia (%)** | Negative | 86 (87.8) | 190 (95.5) | 276 (93.0) |
|  | Positive | 12 (12.2) | 9 (4.5) | 21 (7.1) |
| **Anti-HCMV IgM (%)** | Negative | 92 (93.9) | 186 (93.5) | 278 (93.6) |
|  | Positive | 6 (6.1) | 13 (6.5) | 19 (6.4) |
| **Anti-HCMV IgG avidity (%)** | High-avidity | 97 (99.0) | 198 (99.5) | 295 (99.3) |
|  | Low-avidity | 1 (1.0) | 1 (0.5) | 2 (0.7) |

Data is presented as number and percentage (%) within each category for cases (with tuberculosis) and controls (no tuberculosis), and the combined cohort. CD4 count: among participants with HIV infection only. Rif: rifampicin.

### Association of HCMV DNAemia with tuberculosis disease

The prevalence of HCMV DNAemia was higher among cases than among controls (9, 4.5%; odds ratio [OR] 2.95, 95% confidence interval [95%CI] 1.20–7.47; p=0.019: Table III). After multivariate adjustment for age, HIV status, gender and BMI, this association became stronger (adjusted OR [aOR] 4.99, 95%CI 1.63–16.99; p=0.007). The effect modification analysis showed no significant interaction between HCMV DNAemia and BMI category (interaction term p-values 0.297 and 0.298 for underweight and overweight, respectively; Supplementary table 1).

**Table III.** Logistic regression analysis of factors associating HCMV DNAemia with tuberculosis disease.

| **Factor** | **Category** | **Univariate analysis**<br>**Crude OR (95%CI)** | **P-value** | **Multivariate analysis**<br>**Adjusted OR (95%CI)** | **P-value** |
| --- | --- | --- | --- | --- | --- |
| **HCMV DNAemia** | Negative | 1.00 (Reference) |  | 1.00 (Reference) |  |
|  | Positive | 2.95 (1.20-7.47) | 0.019 | 4.99 (1.63-16.99) | 0.007 |
| **Age** | 18-35 | 1.00 (Reference) |  | 1.00 (Reference) |  |
|  | 36-50 | 0.55 (0.32-0.94) | 0.031 | 0.66 (0.35-1.23) | 0.193 |
|  | >50 | 0.39 (0.19-0.77) | 0.009 | 0.53 (0.24-1.10) | 0.094 |
| **Gender** | Female | 1.00 (Reference) |  | 1.00 (Reference) |  |
|  | Male | 0.78 (0.48-1.26) | 0.308 | 0.47 (0.26-0.83) | 0.011 |
| **HIV status** | Negative | 1.00 (Reference) |  | 1.00 (Reference) |  |
| | Positive (CD4 <200 cells/μl) | 0.25 (0.52-2.88) | 0.605 | 0.64 (0.22-1.75) | 0.391 |
| | Positive (CD4 >200 cells/μl) | 0.90 (0.50-1.59) | 0.726 | 0.61 (0.30-1.18) | 0.149 |
| **BMI** | Healthy weight | 1.00 (Reference) |  | 1.00 (Reference) |  |
|  | Underweight | 2.13 (1.24-3.68) | 0.006 | 2.28 (1.27-4.11) | 0.005 |
|  | Overweight | 0.23 (0.08-0.53) | 0.001 | 0.18 (0.06-0.44) | <0.001 |

OR: Odds Ratio. BMI: Body Mass Index. The three BMI categories have the following ranges; <18.5, 18.5-24.9 and ≥ 25 for Underweight, Healthy weight and Overweight respectively.

### Association of HCMV previous exposure with tuberculosis disease

Comparable proportions between cases and controls were observed in anti-HCMV IgM results with 6.1% (n=6) and 6.5% (n=13) being positive, respectively. Only 2 participants had primary HCMV infection as indicated by low anti-HCMV IgG avidity: 1 case (1.0%) and 1 control (0.5%). Current HCMV reactivation or reinfection was observed in 12 (12.2%) cases and 9 (4.5%) control, recent HCMV reactivation or reinfection in 5 (5.1%) cases and 11 (5.5%) control, and Past HCMV infection in 80 cases (81.6%) and 178 controls (89.4%) (Supplementary table 3).

The number of participants with primary HCMV infection was very small, therefore this exposure category was merged with that of recent HCMV reactivation or reinfection and the combined grouped group as recent HCMV infection, reactivation or reinfection in the subsequent analyses. Participants with current HCMV reactivation or reinfection had significantly higher odds of TB disease compared to those with past HCMV infection (crude OR 2.97, 95%CI 1.21–7.54, p=0.018; aOR 4.88, 95%CI 1.59–16.31, p=0.007) in a multivariable analysis that controlled for age, HIV status, gender and BMI as confounders (Table IV). Tuberculosis disease was not associated with recent HCMV infection, reactivation or reinfection (crude OR = 1.11, 95%CI 0.38–2.97; p=0.837; aOR 1.07, 95%CI 0.33–3.16; p=0.907) in either analysis.

**Table IV.** Logistic regression analysis of factors associating HCMV previous exposure with tuberculosis disease.

| **Factor** | **Category** | **Univariate analysis**<br>**Crude OR**<br>**(95%CI)** | **P-value** | **Multivariable analysis**<br>**Adjusted OR (95%CI)** | **P-value** |
| --- | --- | --- | --- | --- | --- |
| **HCMV**<br>**previous**<br>**exposure** | Past HCMV infection | 1.00 (Reference) |  | 1.00 (Reference) |  |
|  | Current HCMV reactivation or reinfection | 2.97 (1.21-7.54) | 0.018 | 4.88 (1.59-16.31) | 0.007 |
|  | Recent HCMV infection, reactivation or reinfection | 1.11 (0.38-2.97) | 0.837 | 1.07 (0.33-3.16) | 0.907 |
| **Age** | 18-35 | 1.00 (Reference) |  | 1.00 (Reference) |  |
|  | 36-50 | 0.55 (0.32-0.94) | 0.031 | 0.66 (0.35-1.23) | 0.192 |
|  | >50 | 0.39 (0.19-0.77) | 0.009 | 0.53 (0.24-1.10) | 0.095 |
| **Gender** | Female | 1.00 (Reference) |  | 1.00 (Reference) |  |
|  | Male | 0.78 (0.48-1.26) | 0.308 | 0.48 (0.26-0.84) | 0.012 |
| **HIV status** | Negative | 1.00 (Reference) |  | 1.00 (Reference) |  |
| | Positive (CD4 <200 cells/μl) | 0.25 (0.52-2.88) | 0.605 | 0.65 (0.22-1.77) | 0.412 |
| | Positive (CD4 >200 cells/μl) | 0.90 (0.50-1.59) | 0.726 | 0.65 (0.33-1.25) | 0.206 |
| **BMI** | Healthy weight | 1.00 (Reference) |  | 1.00 (Reference) |  |
|  | Underweight | 2.13 (1.24-3.68) | 0.006 | 2.30 (1.29-4.12) | 0.005 |
|  | Overweight | 0.23 (0.08-0.53) | 0.001 | 0.18 (0.06-0.45) | <0.001 |

OR: Odds Ratio. BMI: Body Mass Index. The three BMI categories have the following ranges; <18.5, 18.5-24.9 and ≥ 25 for Underweight, Healthy weight and Overweight respectively.

### Descriptive results of HCMV viral load across related TB disease history/status

The overall range of viral loads was 30.7–16,459.8 and 30.3–46,646.7 copies/ml among cases and controls respectively and the median (IQR) HCMV viral load of the respective groups was 88.9 (47.8–441.3) copies/ml and 107.4 (57.6–153.0) copies/ml. Higher HCMV viral loads were associated with longer time-to-positivity of the mycobacterial culture, i.e. with lower bacterial loads (Supplementary figure 2). Among participants with HIV, cases generally had elevated HCMV viral loads compared to controls (Supplementary figure 3). Furthermore, participants with a history of prior TB demonstrated greater variability in HCMV DNAemia levels, with some individuals exhibiting notably high viral loads, whereas those without prior TB disease mostly had low viral loads (Supplementary figure 4). Cases with previous TB history displayed markedly elevated HCMV viral loads compared to TB cases without past TB history (Supplementary figure 5).

## Discussion

In this nested case-control study, 7.1% of patients diagnosed with TB had active HCMV infection based on HCMV DNAemia. After adjusting for confounding, participants with TB were five times more likely to have active HCMV infection than those without TB. Current HCMV reactivation or reinfection was associated with increased odds of TB disease well as recent HCMV infection, reactivation or reinfection was not.

Our finding that current HCMV infection was strongly associated with tuberculosis in adults aligns with that from a South African longitudinal birth cohort study(5). Unlike the Martinez *et al*. study which reported that active HCMV replication in infancy predicted later TB disease, our cross-sectional study explored the concurrent relationship between HCMV and TB among adults. Our findings show that current HCMV infection and TB are associated when measured at the same time point but showed no association of active TB with earlier HCMV exposure (recent HCMV infection, reactivation or reinfection). These findings suggest that while primary HCMV infection may influence TB susceptibility early in life, recent HCMV infection, reactivation or reinfection plays no or only a limited role in adult TB pathogenesis. The childhood and adulthood tuberculosis risk attributed to HCMV may be different because unlike adults in TB endemic countries who are likely to experience reactivation or reinfection with HCMV and Mtb, children are more likely to have primary diseases due to both pathogens.

HCMV repeatedly infects immunocompetent individuals(15-17) and these infections become persistent and incremental leading to superinfections with same or different genotypes in humans(18). Persistent infection with HCMV leads to immunosenescence through immune inflation where there is depletion of naïve T cells and a tremendous expansion of effector memory CD8^+^ T and CD4^+^ T cells(19) due to chronic immune activation. Immune activation due to HCMV determined by IFNγ ELISPOT positivity in South African children was associated with a moderately high TB disease risk(9). Children are expected to have less immune activation due to HCMV compared to adults who experience incremental exposure especially in endemic areas. A study done among Ugandan adults(20) associated HCMV specific IgG titers and inflammation which was measured by IP-10 and IL-1α with increased TB disease risk. HCMV reactivation or reinfection however may have a smaller role in immune activation in adults which may be due to chronic infections other than HCMV(21), age-related immunosenescence (22), and environmental agents and lifestyle factors(23), amongst others.

The observed association of current HCMV reactivation or reinfection but not recent HCMV infection, reactivation or reinfection with TB as observed in this study may suggest that it is TB disease leading to reactivation of HCMV. Indeed, HCMV reactivation may be a consequence of immune activation because the virus is mostly detected from tissues undergoing inflammation(24) and TB disease is characterized by extensive inflammation. Inflammation due to TB disease may lead to development and multiplication of cells containing HCMV and ultimately lead to its replication as a consequence of IFN-γ and TNF-α cytokines produced by activated T cells(25). An alternative hypothesis may be that underlying decline in cellular immune functionality with age(26) which may trigger both progression to TB disease and HCMV reactivation.

For the descriptive findings, higher HCMV viral loads were associated with longer mycobacterial culture time-to-positivity which may suggest that viral activity may correlate with lower bacterial burden. This contrasts with previous studies reporting a dose-response relationship between CMV exposure and TB risk(5, 20). Among participants with HIV, cases generally had elevated HCMV viral loads compared to controls, consistent with studies that have found impaired immune control of HCMV and increased viremia in people living with HIV(27, 28). The increased viral load in cases could be due to increased inflammatory environment in cases than in controls. Individuals with prior TB showed greater variability in HCMV DNAemia, with some cases exhibiting markedly high viral loads, possibly reflecting episodes of viral reactivation triggered by immune modulations. These observations suggest that HCMV reactivation dynamics may be influenced by both host immune status and TB disease history.

Our study had several limitations that could affect interpretation and generalization of the findings. Due to the cross-sectional design, we could not exclude with certainty that HCMV exposure preceded TB disease. We could not control for potential residual confounders like immunological status (beyond HIV), socio-economic status, behavioral factors such as alcohol use, co-infections, comorbidities, and host genetics. The Euroimmun HCMV specific IgM ELISA kits were reported to have a low sensitivity for serum samples stored beyond 7 weeks(29) and hence we could have misclassified HCMV exposure categories. The high proportion of individuals with history of previous TB treatment and those who are of mixed-race ancestry may affect the generalization of our results to other settings.

In conclusion, our findings show that TB disease was strongly associated with current active HCMV infection but not with recent HCMV infection, reactivation or reinfection. This may indicate that among adults in high HCMV and TB incidence settings, active TB disease reactivates HCMV replication rather than the other way around. Prospective studies carried out in multiple settings are needed to determine the direction of causality as well as accounting for residual confounders.

## Data Availability

All data generated or analyzed during this study are included in this published article and its supplementary information files

## Author contributions

DS, AG-B, MLJ, FC and WS conceptualized the idea. AC, BWPR, ZP, HM and RW collect samples and collated data. DS, AG-B, FC, GT and WS performed the statistical analyses. DS drafted the first manuscript. All authors edited and reviewed the manuscript.

## Acknowledgment

The authors would like to thank BAR-TB Dx participants and the study team for their contribution to this study. FC received support from EDCTP and Mr. Willem Bakhuys Roozeboomstichting well as DS received the co-funding from SANTHE.

